# Parental attitudes towards mandatory vaccination; a systematic review

**DOI:** 10.1101/2021.02.24.21250288

**Authors:** Louise E. Smith, Ava Hodson, G. James Rubin

## Abstract

While mandatory vaccination schemes can increase vaccine uptake rates, they can also cause backlash among some parents. We conducted a systematic review investigating parental beliefs about vaccine mandates and factors associated with support for mandatory vaccination schemes. We searched Embase, Ovid MEDLINE, Global Health, APA PsycINFO and Web of Science from inception to 17^th^ September 2020. Seventeen studies (five qualitative, twelve quantitative) were eligible for inclusion. We synthesised results of qualitative and quantitative studies separately. Studies were heterogeneous with regard to schemes investigated and factors investigated. Quantitative studies found little evidence for any factors being consistently associated with support for mandatory vaccination. Qualitative studies found that parents perceived mandatory vaccination schemes as an infringement of their rights and that they preferred universal, compared to targeted, schemes. To optimise engagement with existing child mandatory vaccination legislation, schemes should be designed with parental beliefs in mind.

## Introduction

While vaccines have substantially reduced morbidity and mortality for various diseases,[1] some parents choose not to vaccinate their child. In the UK, uptake of routine childhood vaccines has been decreasing for the last five years.[2] Vaccine uptake has also been decreasing in the United States (US) and elsewhere globally.[3] One way of increasing uptake is make childhood vaccination mandatory. As of December 2018, 105 countries had a nationwide vaccine mandate in operation.[4] Other countries such as the US and Canada, do not have a nationwide mandate, but have mandates that vary on a regional basis.[5-7] Mandatory vaccine schemes tend to restrict access to child-care or schooling for children who are not vaccinated, or withhold state payments or benefits if children are not vaccinated.[4] In some circumstances, parents can apply for exemptions to vaccine mandates based on religious or personal beliefs, but the flexibility within policies varies widely.[4] Another way of promoting vaccination could be to offer financial incentives.

Making vaccinations mandatory tends to increase uptake.[8] However, there is substantial debate over the ethics of making vaccination mandatory.[9] While mandatory vaccination may force people to overcome barriers to vaccination, such as having to make a primary care appointment, it may entrench negative perceptions of vaccination.[10] In some countries, the implementation of mandatory vaccination programs has led to increased anti-vaccination sentiments and negative vaccine messages in the media.[11, 12] In Germany, these negative sentiments impacted vaccination intentions for other recommended, but non-mandatory, vaccinations.[12] In the US, systematic differences in uptake of child vaccines still exists, with evidence of geographical clustering of vaccine exemptions.[13]

There is no standard approach to mandatory vaccination programmes.[14] Approaches vary country to country by: which vaccines are mandatory; which age groups are included; and how flexible the mandate is (e.g. penalties, enforcement, ability to opt out, compensation for serious adverse events). Refusal is often allowed based on religious, moral, philosophical or personal reasons. To minimise backlash when implementing a new childhood vaccine mandate, and to optimise engagement with existing childhood mandatory vaccination legislation, schemes should be designed with parental beliefs in mind. One recent systematic review investigated parental beliefs towards mandatory vaccination, h but with major limitations.[15, 16] Another systematic review focused on acceptability, economic costs and incentives of specific schemes, but is now outdated having been conducted in 2013.[17] There are no recent, high quality reviews investigating parental beliefs and attitudes towards vaccine mandates for routine childhood vaccinations.

The aim of this study was to investigate parents’ beliefs about vaccine mandates and factors associated with support for mandatory vaccination schemes.

## Method

We conducted a systematic review in accordance with PRISMA criteria [18] to investigate parents’ beliefs and attitudes towards vaccine mandates and mandatory vaccination. We searched Embase, Ovid MEDLINE, Global Health and APA PsycINFO through OvidSP, and Web of Science. Our final search term combined terms related to: mandatory, compulsory, exemptions, or school entry requirements; vaccination or immunisation; beliefs, or attitudes; and children (see supplementary materials). Databases were searched from inception to 17^th^ September 2020. References and forward citations of included articles were also searched.

### Inclusion criteria

The following inclusion criteria were used:

#### Participants

Studies were included if they investigated parental beliefs. Studies were excluded if they investigated healthcare workers’ beliefs about vaccine mandates, or if they investigated attitudes towards mandatory vaccination in populations other than children.

#### Predictors/exposures

Studies were included if they investigated factors associated with vaccine mandates or support for mandatory vaccination schemes, or if endorsement of vaccine mandates or mandatory vaccination was described.

#### Outcome

Beliefs and attitudes about vaccine mandates for routine childhood vaccinations or the concept of mandatory vaccination. Studies were only included if they investigated a WHO recommended routine vaccination in children (BCG, hepatitis B, polio, DTP-containing vaccine, haemophilus influenzae type b (Hib), pneumococcal (conjugate), rotavirus, measles, rubella).[19] We excluded studies investigating beliefs about mandates for HPV vaccination, as parental concerns may be influenced by complex underlying attitudes towards sex,[20] which are likely qualitatively different to concerns surrounding vaccine mandates for other routine childhood vaccinations. Studies were also excluded if they solely investigated factors associated with uptake of mandatory vaccines.

#### Study reporting

Studies were included if they were published in English and presented data. There were no exclusions made based on study design.

### Data extraction

We extracted information about study design, inclusion criteria, participant characteristics, country of study, and attitudes towards vaccine mandates or mandatory vaccination.

### Risk of bias

Risk of bias was measured using the mixed methods appraisal tool (MMAT).[21] This tool allows appraisal of different study methods, including qualitative and quantitative studies. The MMAT evaluates studies on five dimensions which vary based on the methods of the study. To enable more detailed description of risk of bias in the study, we scored studies out of two on each of the five dimensions, resulting in a total score out of ten. To aid interpretation of results, studies were rated as poor quality if they scored five or under; moderate quality if they scored six or seven; and good quality if they scored eight or over.

LS and AH completed risk of bias ratings separately for all studies. Any discrepancies were solved through discussion and final scores were approved by both authors.

### Procedure

LS came up with the search terms, carried out the search, screened papers, extracted data and completed risk of bias assessment. AH screened a random sample of 100 citations to full-text screening stage and completed risk of bias assessment. Guidance was provided by GJR. Qualitative and quantitative data were synthesised separately. Quantitative data were narratively synthesised, considering risk of bias ratings. As studies were heterogeneous in the mandatory vaccination schemes and associated factors investigated, there was no scope to conduct a meta-analysis. Qualitative data were synthesised using meta-ethnography,[22] synthesising themes reported across studies included.

## Results

### Study characteristics

The search identified 4,994 citations; after removing duplicates, 2,672 citations remained. After title, abstract and full-text screening, seventeen citations remained. A further three citations were identified by reference searching and forward citation tracking. Thus, twenty citations, reporting on seventeen studies (twelve quantitative, five qualitative) met inclusion criteria (see Figure 1).

**Figure 1.**
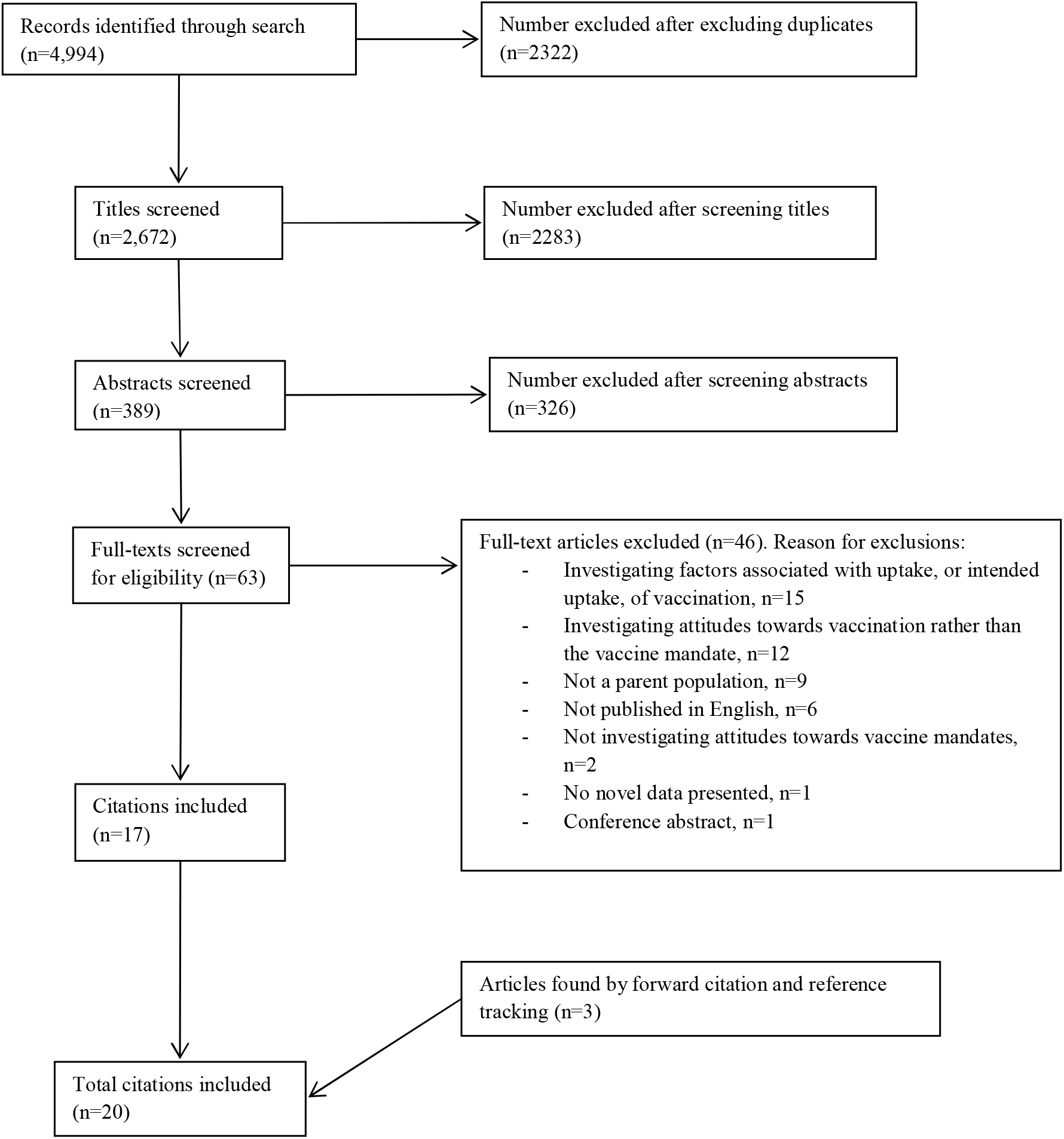
Flowchart depicting the selection of studies for the systematic review, with reasons for exclusion

### Risk of bias

Scores for studies ranged between three and ten (see supplementary materials). Qualitative studies scored highly, between nine and ten,[17, 23-27] aside from one short article which scored poorly (five).[28] Two quantitative studies scored highly (eight or nine),[29, 30] and were only marked down for not reporting complete outcome data (including 95% confidence intervals for prevalence estimates) or fully accounting for confounders in the study design or analyses. Five quantitative studies scored six or seven.[17, 23, 31-35] These studies did not report complete outcome data, account for confounders in the study design or analysis, and participants were not representative of the target population or the outcome measure was not robust. Five quantitative studies scored particularly poorly (between three and five).[36-41] These studies were poor across the different dimensions evaluated, but low scores were because participants were not representative of the target population, complete outcome data were not reported, confounders were not accounted for in the study design and analysis, and outcome variables were not robustly measured.

### Quantitative studies

Ten studies used a cross-sectional design, one used a case-control design, and one study was a discrete choice experiment (see supplementary materials). Studies were conducted in the USA (n=5), Poland (n=2), Israel (n=2), England (n=1), Croatia (n=1), and Australia (n=1). Studies investigated factors associated with support for various mandatory vaccination schemes, including: restricting access to childcare or schooling for children who are not vaccinated; removing religious and personal belief exemptions from childcare or school entry requirements; withholding state payments or benefits if children are not vaccinated; and mandatory vaccination generally.

Support for mandatory vaccination schemes varied. One study found that 47% of rural Ohio Appalachian (US) parents who had not vaccinated their daughters for HPV believed they had the right to refuse vaccines that were require for their child’s school,[35] while another found that 12% of US parents believed that children should be allowed to go to school even if they were not vaccinated.[30] Support for religious belief exemptions (22%) was slightly higher than support for personal belief exemptions (17%) in US parents.[34] Another study found that 73% of Croatian parents of children aged 6 years, attending school health services believed that child vaccination should remain mandatory.[39] In a sample of Australian parents of children under 5 years, 82% supported the “no jab, no pay” scheme.[29]

Four studies found no evidence for an association between parent gender and support for mandatory vaccination schemes.[29, 30, 35-37] One poor quality study found an association between younger age and lower support for mandatory vaccination,[36, 37] while three studies (two good quality, one moderate quality) found no evidence for an association.[29, 30, 35] A further poor quality study found that parents of younger children were less likely to support mandatory vaccination.[41] Lower education was associated with lower support for mandatory vaccination in a moderate quality study,[32] but not a poor quality study.[36, 37] Two further studies (one good quality, one moderate quality) found no evidence for an association between education and support for mandatory vaccination schemes restricting access to child-care or schooling only to vaccinated children.[30, 35] Lower household income was associated with lower support for mandatory vaccination schemes in which childcare or schooling is limited to vaccinated children in a good quality study,[30] while a moderate quality study found no evidence for an association.[35] Lower household income was also associated with lower support for mandatory vaccination in a poor quality study.[36, 37] White ethnicity was associated with support for restrictions in eligibility for childcare or schooling for unvaccinated children.[30] There was no evidence for an association between ethnicity and the “no jab, no pay” scheme.[29] Both studies investigating associations with ethnicity were good quality studies. Larger household size [30] and not being in full-time employment [35] were associated with lower support for a school-entry or childcare entry-based vaccination scheme. Poorer health literacy was associated with lower support for mandatory vaccination in one moderate quality study.[32] There was no evidence for an association between the child having previously experienced an adverse effect from vaccination and support for mandatory vaccination in a poor quality study.[36, 37] There was also no evidence that support for mandatory vaccination schemes changed between 2008 and 2016 in a moderate quality study.[33]

Parents of US children who were home-schooled were less supportive of mandatory vaccination,[40] and more supportive of allowing religious belief exemptions and personal belief exemptions for vaccines required for school-entry.[34, 40] In an Australian study, parents who had previously registered a conscientious objection to vaccination were less likely to support the “no jab, no pay” scheme.[29] Parents in a US study who lived in a state which allowed philosophical exemptions were less likely to support mandatory vaccination for childcare or schooling.[30]

Distrust of the government and of healthcare providers was associated with lower support for mandatory vaccination for childcare or schooling,[38] as was Republican and Independent political affiliation (compared to Democrat) in one US study.[35]

One Australian study found that parents who perceived a lower risk of measles for unvaccinated children were less likely to support the “no jab, no pay” scheme; there was no evidence for an association with needing the financial incentives to afford family expenses or having a child that attends childcare.[29]

### Qualitative studies

Two studies, both conducted in the UK, used a focus group design. Three other studies (conducted in Australia, the USA and Hong Kong) used an interview design. Studies investigated parental beliefs about mandatory vaccination schemes which: offer financial incentives for vaccination; restrict access to childcare or schooling for children who are not vaccinated; remove religious and personal belief exemptions from childcare or school entry requirements; withhold state payments or benefits if children are not vaccinated; and which restrict access to child-care or schooling and withhold state payments or benefits if children are not vaccinated.

Seven main themes were extracted from studies (see Table 1). First, parents consistently perceived mandating vaccination as an infringement of their personal rights.[17, 23, 25, 26, 28] In a sample of parents who had refused mandatory vaccination, the mandate had strengthened their commitment to make autonomous medical decisions for themselves and their child.[24] Second, parents thought that schemes should not “punish” the child by not allowing them access to schooling or childcare based on the parents’ choice not to vaccinate them (parental preferences for mandatory vaccination schemes).[17, 23, 24, 26] However, they perceived restricting access to schooling and childcare as fairer and more equitable than offering financial incentives for vaccination, and preferred universal rather than targeted schemes.[17, 23, 26] Third, parents perceived financial incentives for vaccination as inappropriate and coercive,[17, 23, 24, 26] and thought that parents may use vaccination schemes as a way to gain additional financial incentives or to secure school spaces (perceived inappropriateness of schemes).[28] Fourth, motivators for vaccination were varied. While one study found that parents thought that protecting the child should be the sole motivation for vaccination,[28] other studies found that parents thought mandatory vaccination could be an incentive for vaccination if their child would otherwise be denied schooling,[27] or if financial incentives could be seen as supplementing one’s income.[17, 23, 26] Fifth, mandatory vaccination schemes were perceived as having a disproportionate impact on low-income families, who may be more reliant on state benefits, financial incentives and who might not have the resources to pay for alternate schooling or childcare.[17, 23, 26, 28] Sixth, parents objected to penalising parents who did not want to vaccinate their child due to safety concerns.[28] Seventh, parents agreed that mandatory vaccination would deliver some “peace of mind” through the knowledge that there was no risk of unvaccinated children passing on illnesses in school or childcare settings.[17, 23, 26, 27]

**Table 1.** Themes and sub-themes identified in studies included

| Theme | Sub-themes |
| --- | --- |
| Infringement of parental rights | - Infringement of parental rights and autonomy [17, 23, 26]; [25]<br>- Disempowerment of parents by removal of consent [28]<br>- Commitment to autonomy strengthened [24] |
| Parental preferences for mandatory vaccination schemes | - Mandatory vaccination schemes should not “punish” the child (e.g. by not being able to go to school) rather than the parent [17, 23, 26]; [24]<br>- Restricting access to child-care and schooling only to vaccinated children is perceived as being preferable, fairer and more equitable to offering financial incentives for vaccination [17, 23, 26]<br>- Universal schemes seen as more equitable and fairer than targeted schemes (e.g. for those with unvaccinated children, lower socio-economic status). [17, 23, 26]<br>- Disdain towards withholding state benefits and payments if children are not vaccinated [24] |
| Perceived inappropriateness of schemes | - Financial incentives seen as inappropriate and coercive [17, 23, 26]; [24]<br>- Schemes requiring vaccination for child-care or schooling and withholding state benefits inappropriately position vaccination as a means to acquire financial benefits and secure a school space [28] |
| Motivation for vaccination | - Protecting child from illness should be sole motivation in parents’ vaccination decision [28]<br>- If perceived importance of schooling is high, making vaccination mandatory for child-care and schooling may act as a driver for vaccination [27]<br>- Mandatory vaccination schemes could facilitate normalisation of vaccination behaviour and encourage those who do not prioritise vaccination [17, 23, 26]<br>- Parents who did not vaccinate their child reported that no government measure would change parents’ minds about vaccinating their child [24]<br>- Introduction of mandatory vaccination schemes may be seen positively in disadvantaged groups as ways of supplementing income [17, 23, 26] |
| Disproportionate impact on low-income families | - Incentives, penalties and pressure to vaccinate may be more keenly felt by parents from low-income families who may be more reliant on financial incentives of vaccination, state benefits, and who would not have the financial resources to pay for alternative child-care or schooling if their child could not attend publicly-funded schooling [28]; [17, 23, 26]<br>- May create greater divide between parents who can and cannot afford to vaccinate their child [17, 23, 26] |
| Safety concerns | - Objection to penalising parents who did not want to vaccinate their child due to concerns about vaccination safety [28]<br>- No fault vaccine injury compensation scheme in “No jab, no Pay” [24] |
| Risk of unvaccinated children in passing on illnesses | - Agreement that unimmunised children should be excluded from interacting with other children in child-care or school settings [17, 23, 26];[27]<br>- Give parents “peace of mind” [17, 23, 26] |

## Discussion

Dropping vaccination rates, such as those seen in the US, UK, and elsewhere,[2] and the recent COVID-19 pandemic have re-ignited discussion about mandatory vaccination.[42] While introducing vaccine mandates increases uptake of vaccines,[8] they also have the potential to cause backlash. This may be particularly true for vaccine mandates that are considered to be more stringent.[4] When implementing mandatory childhood vaccination, parents’ viewpoints should be considered, to ensure schemes are accepted and to minimise backlash. This review synthesised quantitative and qualitative studies investigating parental beliefs about mandatory vaccination and factors associated with support for vaccine mandates.

Perhaps unsurprisingly, all but one qualitative study found that parents thought schemes were an infringement of their right to choose whether to vaccinate their child. In a sample of parents who had not vaccinated their child, the introduction of a scheme withholding state benefits if children were not vaccinated strengthened their commitment to autonomy in making health decisions.[24] Recent evidence shows that attitudes towards vaccinations are becoming more polarised.[43] Another theme identified in this study was that parents thought they should not be penalised for not wanting to vaccinate their child due to safety concerns. A quantitative study found that support for religious belief exemptions (22%) was slightly higher than support for personal belief exemptions (17%) in US parents.[34] Implementing vaccine mandates without considering parents’ views on exemption policies could result in considerable backlash among some parents, however the purposive use of non-representative samples in the qualitative studies makes it impossible to identify the prevalence of these views.

Better insight into how many parents this might apply to can be found in quantitative studies. These indicated that support for mandatory vaccination schemes in countries where one had been implemented was reasonably high, ranging between 73% and 88% of parents. Support for a mandatory vaccination scheme was much lower in one study (47%),[35] although this was in a sample of parents who had not vaccinated their daughters for HPV.

Studies investigated support for different vaccine mandates in which unvaccinated children could not access schooling or childcare, parents did not receive state benefits, or in which parents received financial incentives, and for different aspects of vaccine mandates, such as support for specific exemption schemes. Due to the lack of a standard approach to mandatory vaccination,[14] it is difficult to quantify support for mandatory schemes generally. A more useful approach may be to identify parental preferences for different vaccination schemes. One study included in the review did this, finding that parents preferred universal mandatory vaccination schemes compared to those which targeted parents who had not fully vaccinated their child.[17, 23, 31] Qualitative results also indicated that universal schemes were perceived as fairer and more equitable than targeted schemes,[17, 23, 26] and that schemes offering financial incentives for vaccination are perceived by some as inappropriate and coercive. [17, 23, 24, 26] Where financial incentives were offered, cash incentives were preferred by parents of children “at risk” for incomplete vaccination (parents of children aged under 5 years currently living in a deprived area; who were aged 20 years or under; single parents or guardians; who have more than three children; and who had a child aged under 5 years with a physical or mental health condition) compared to no incentive, with no preference for voucher incentives compared to no incentives.[17, 23, 31] There was no evidence for this preference in parents of children “not at risk” (parents of children aged under 5 years who did not fit into the previously listed categories).

Despite ethical concerns about mandating vaccinations,[9] another theme extracted from qualitative studies was the view among some parents that unvaccinated children should be excluded from interacting with other children at school or in childcare settings to reduce the risk of passing on illnesses. This would give parents “peace of mind” about the safety of their child.

Due to heterogeneity of the quantitative studies, there was little evidence for associations between parental support for mandatory vaccination schemes and socio-demographic characteristics and beliefs. There was some evidence that lower household income was associated with lower support for mandatory vaccination schemes. This may reflect lower patterns of vaccine uptake in children of parents with lower incomes generally.[44] One theme extracted from qualitative studies was that mandatory vaccination schemes may have a disproportionate impact on low-income families, with parents being less financially able to seek alternative childcare or schooling arrangements if their child is not vaccinated, or being more reliant on state benefits or financial incentives only given to those whose children are vaccinated. Qualitative findings suggested that disadvantaged groups might support financial incentives as a way of supplementing income.[17, 23, 26] However, there was no association between support for a scheme withholding state benefits for parents of unvaccinated children and needing the benefits to be able to afford family expenses or having a child that attended childcare.[29] Mandatory vaccination schemes should aim to minimise disproportionate impact on low-income families, so as not to create further inequity.

### Limitations of studies included in the review

While qualitative studies included in the review were generally of high quality, quantitative studies were of lower quality. In particular, quantitative studies often did not ensure their samples were representative of the target population or take potential confounders into account in the study design or analyses. Furthermore, outcome data was often not completely reported, and some studies used outcome measures which were not methodologically robust. Some studies investigating parental beliefs about mandatory vaccinations were conducted in countries where vaccination was not mandatory. While this is an important first step if new mandatory vaccination schemes are to be implemented successfully, intentions and concerns about hypothetical situations may not reflect real-world experiences.

### Limitations of the review

Studies were heterogeneous in terms of mandatory vaccination schemes investigated. Parental concerns and beliefs about schemes mandating vaccination for schooling or childcare may be qualitatively different from schemes using financial incentives to promote vaccination. Few quantitative studies investigated the same predictive factors. This lack of replication meant that even where multiple studies found an association, evidence for that factor remained weak. We have based our interpretation and conclusions on factors that were supported by evidence from both quantitative and qualitative studies.

MeSH terms were not searched, therefore some studies which were eligible for inclusion could have been missed.

## Conclusion

While mandatory vaccination schemes increase vaccine uptake rates, they have the potential to cause backlash in some parents. Results from qualitative studies indicated that mandatory vaccination schemes were perceived by some parents as an infringement of their rights. Nevertheless, some parents also felt that schemes limiting access to schooling of unvaccinated children gave them “peace of mind.” Parents preferred universal vaccination schemes, rather than targeted schemes, and particularly disliked schemes offering financial incentives for vaccination. However, these results should be interpreted with caution, taking into account the purposive use of non-representative samples. Quantitative studies reporting rates of endorsement of these views found that support for mandatory vaccination schemes was reasonably high (73% to 88%). Due to heterogeneity of quantitative studies, there was little evidence for factors consistently associated with support for mandatory vaccination. Parental beliefs about vaccine mandates and factors associated with support for mandatory vaccination may shed light on how to implement schemes to maximise parental endorsement.

## Data Availability

N/A

## Supplementary materials. Full details of search

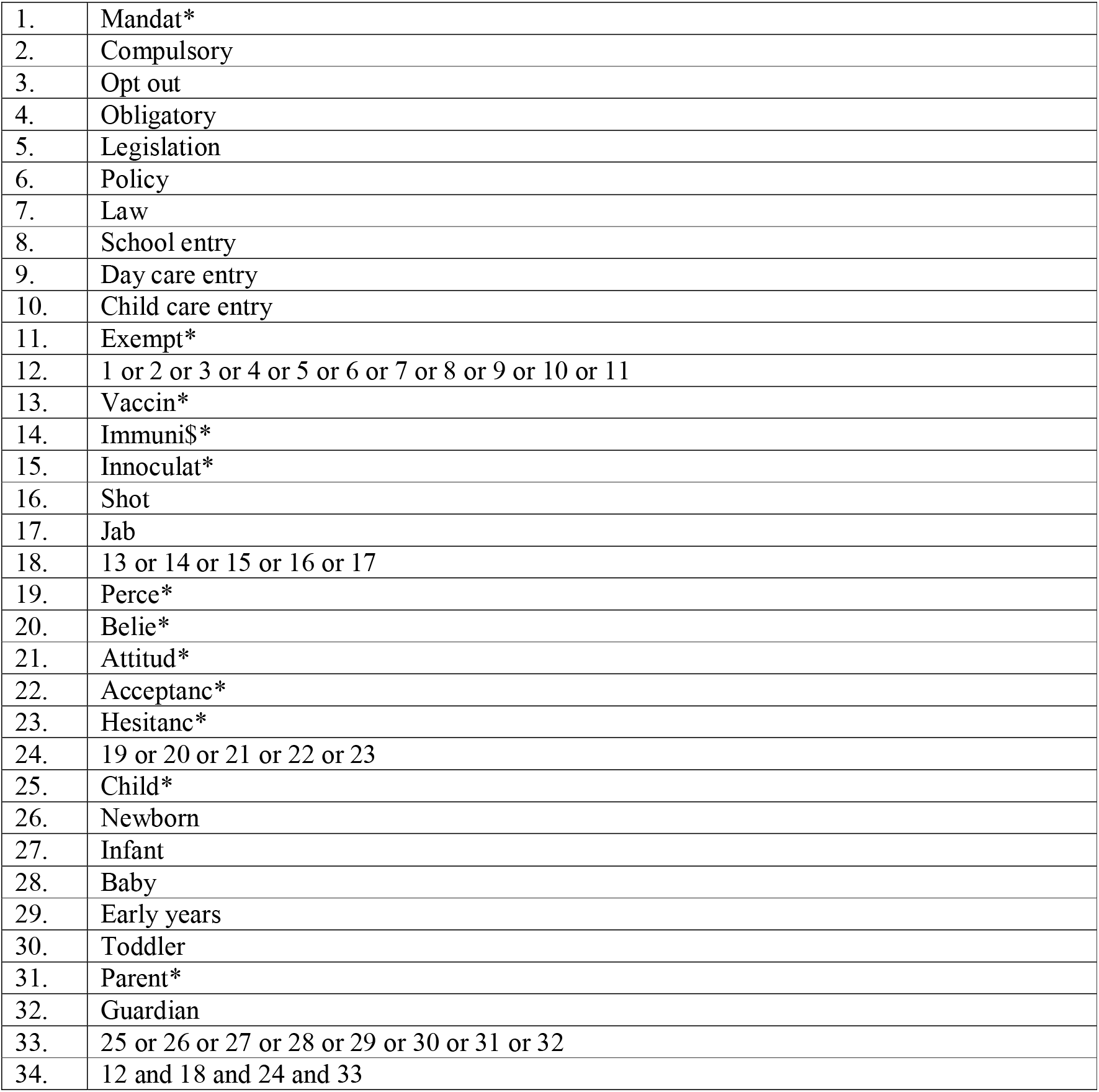

Databases searched:

- Embase: 1974 to 2020 week 37
- Ovid MELINE ® and Epub Ahead of Print, In-Process & Other Non-Indexed citations and daily: 1946 to September 16, 2020
- Global Health: 1973 to 2020 Week 37
- APA PsycInfo: 1806 to September week 1 2020
- Scopus: 1788 to September 17 2020
- Web of Science core collection: data last updated 2020-08-19
  - Science Citation Index Expanded (SCI-EXPANDED) --1900-present
  - Social Sciences Citation Index (SSCI) --1956-present
  - Arts & Humanities Citation Index (A&HCI) --1975-present
  - Conference Proceedings Citation Index-Science (CPCI-S) --1990-present
  - Conference Proceedings Citation Index-Social Science & Humanities (CPCI-SSH) --1990-present
  - Emerging Sources Citation Index (ESCI) --2015-present

## Supplementary materials. Full table of methods and results of studies included in systematic review

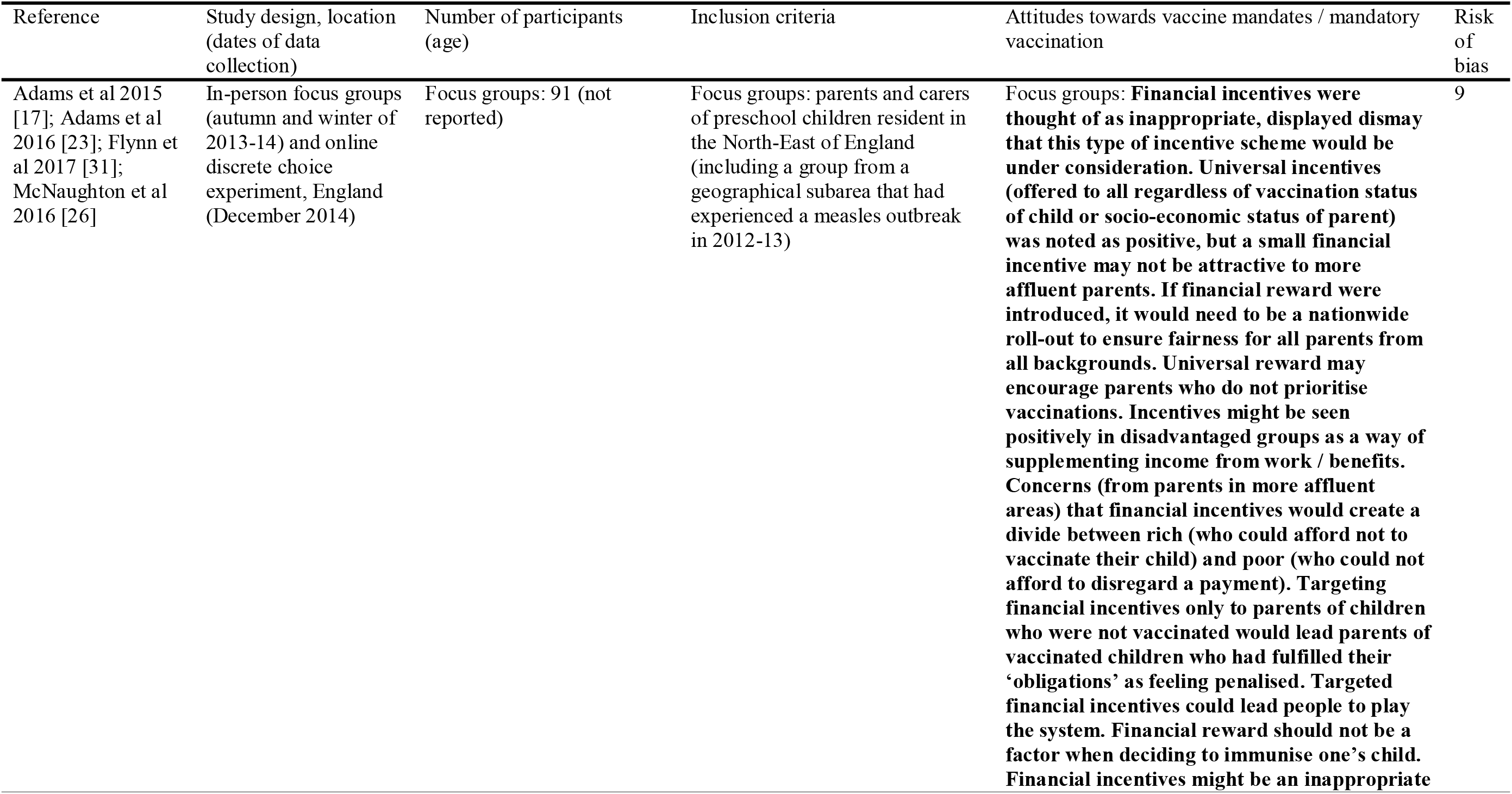
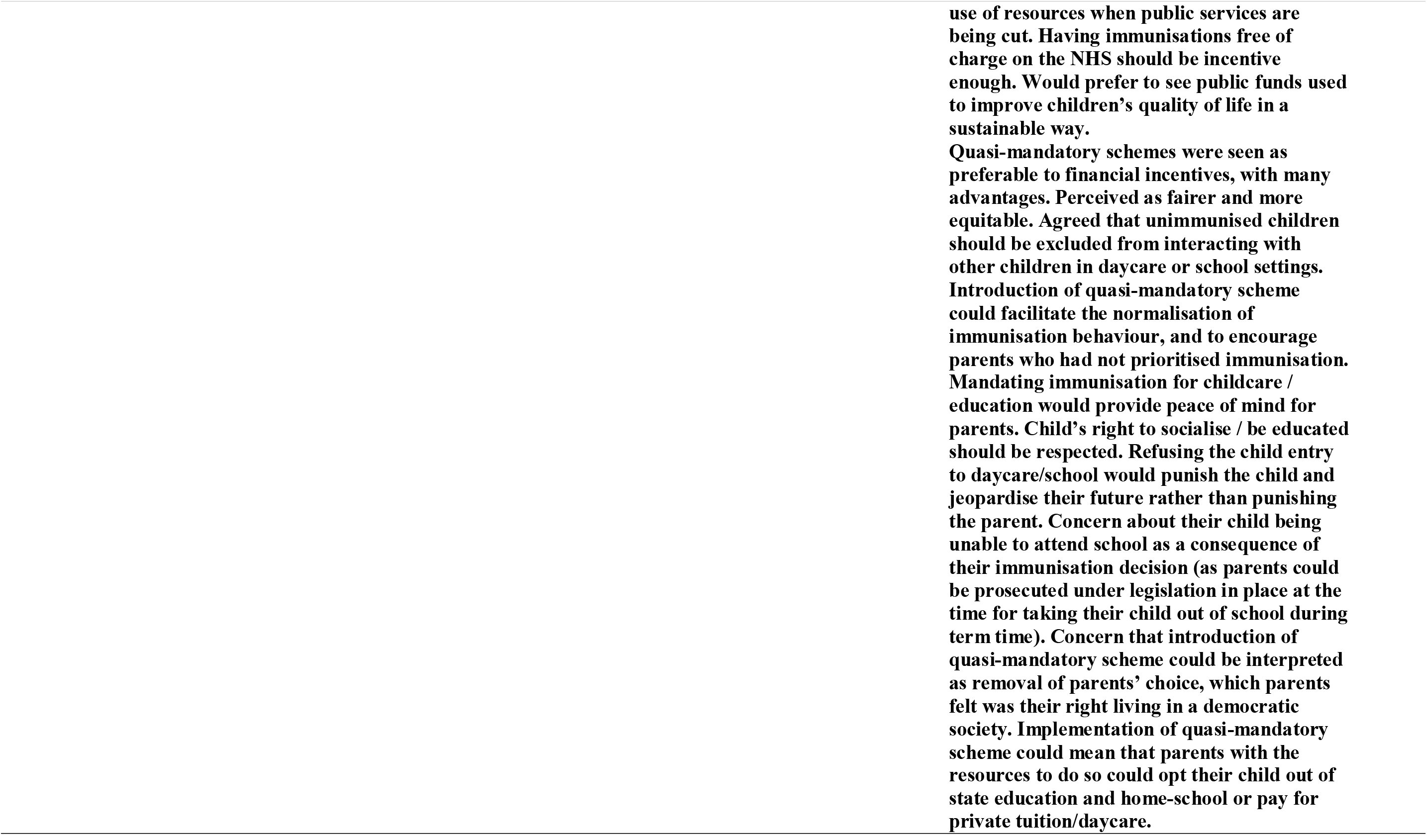
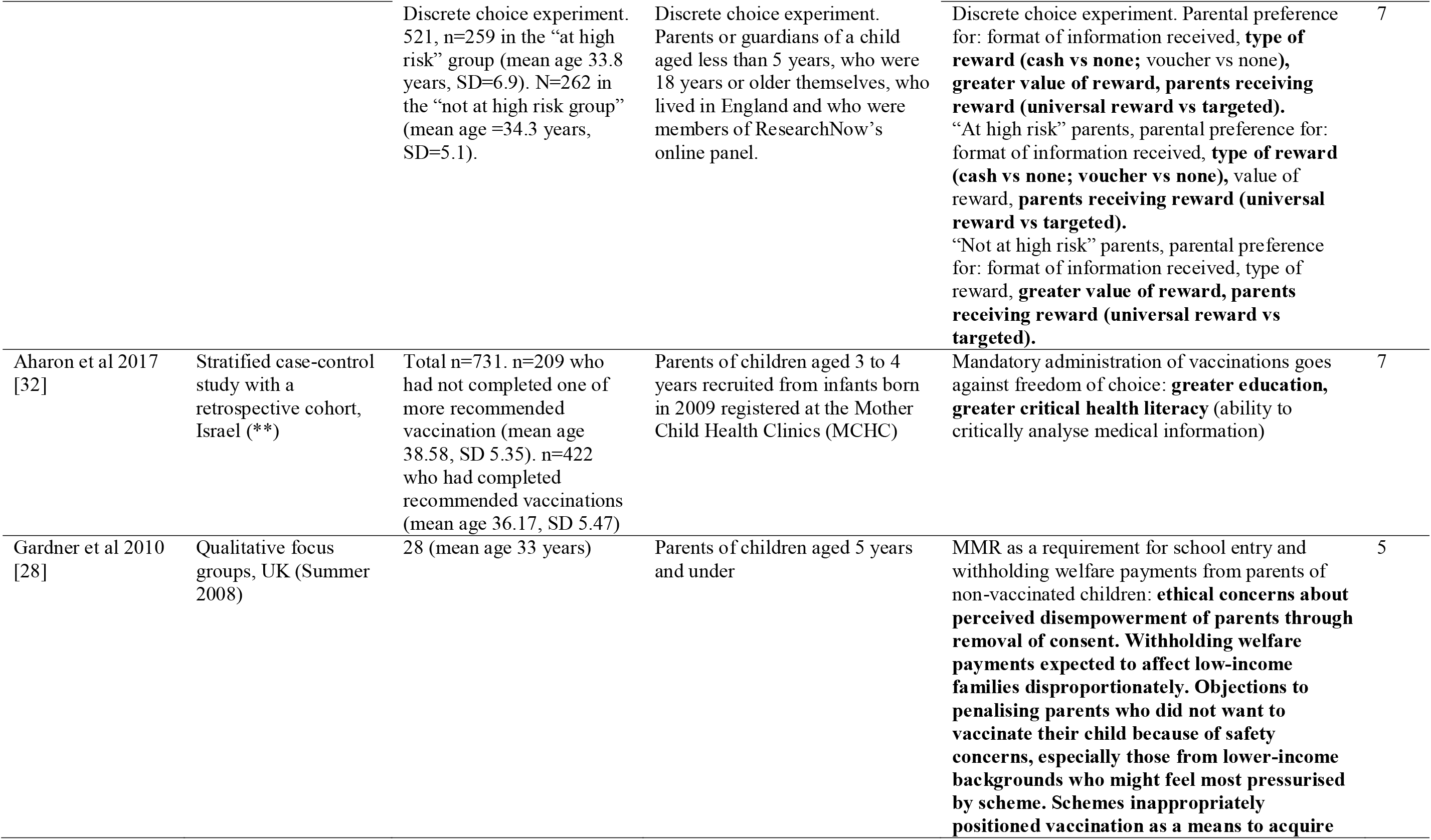
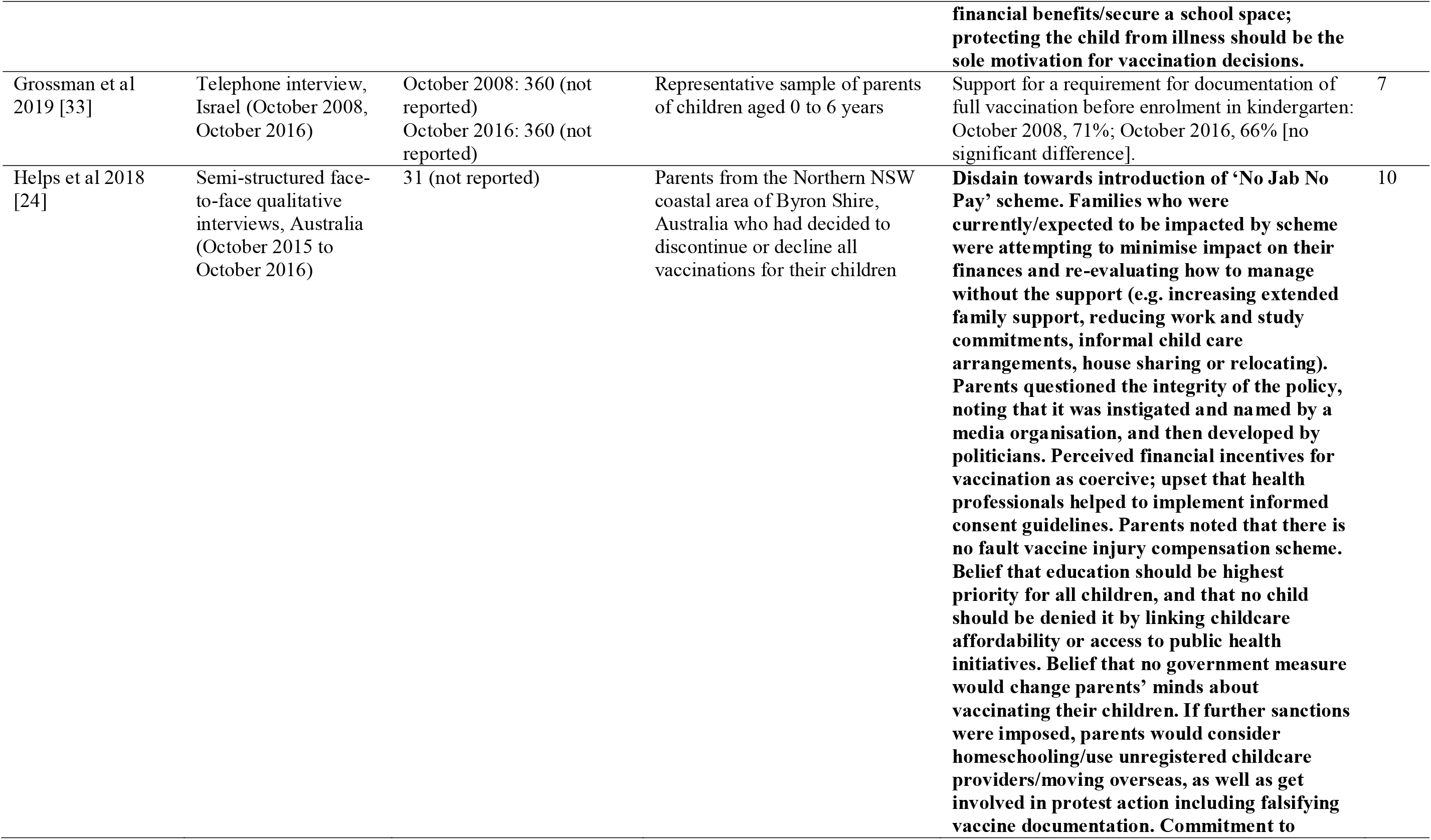
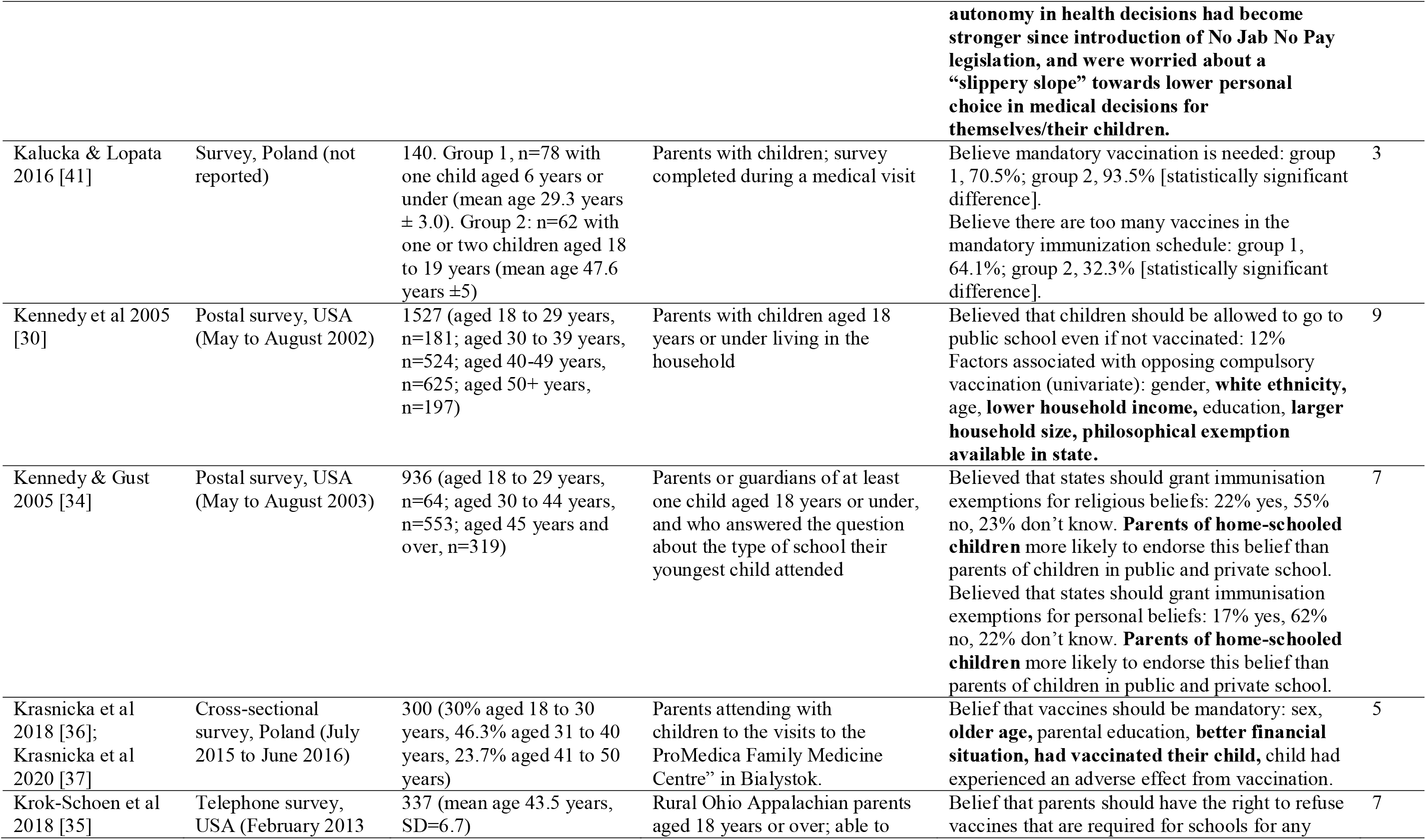
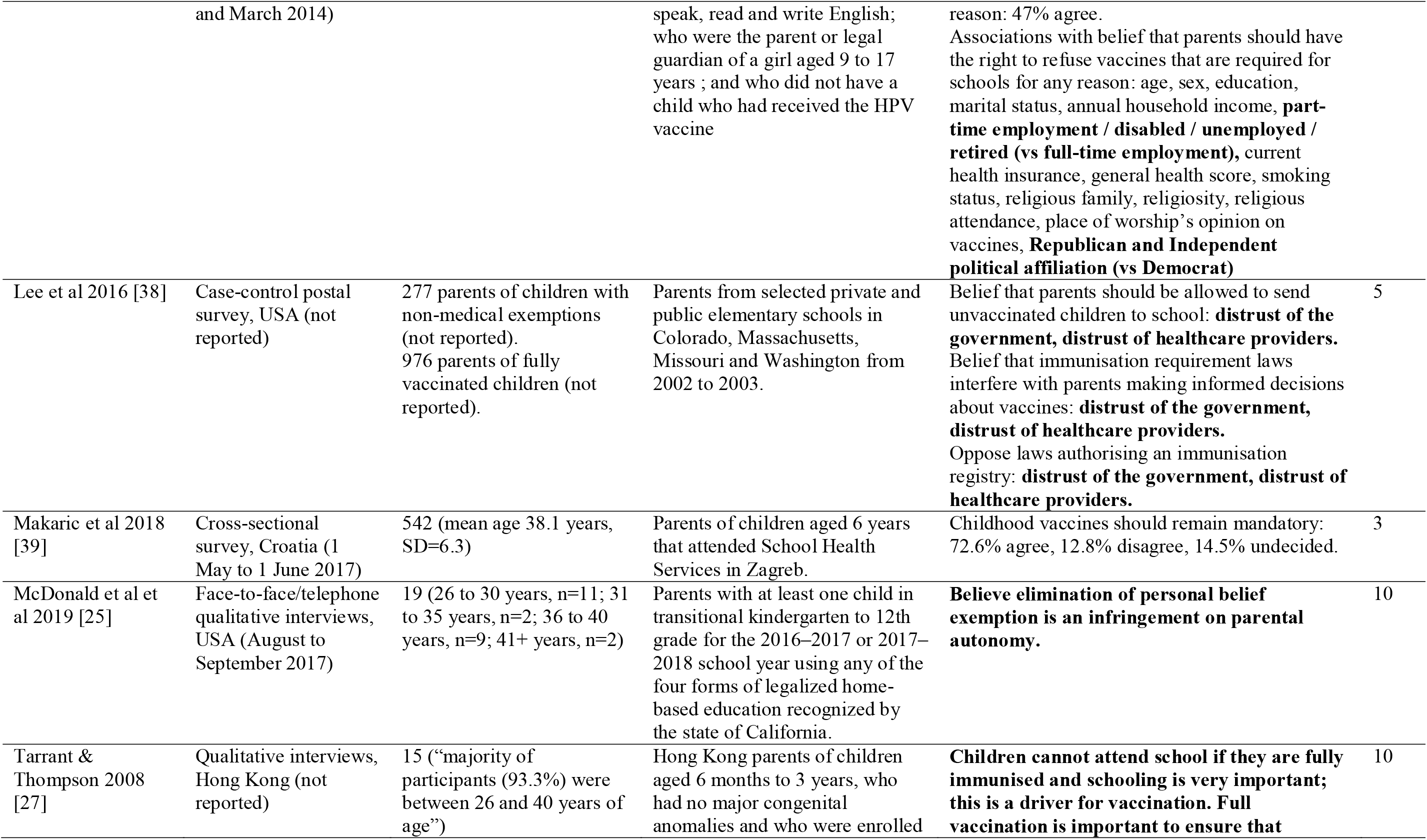
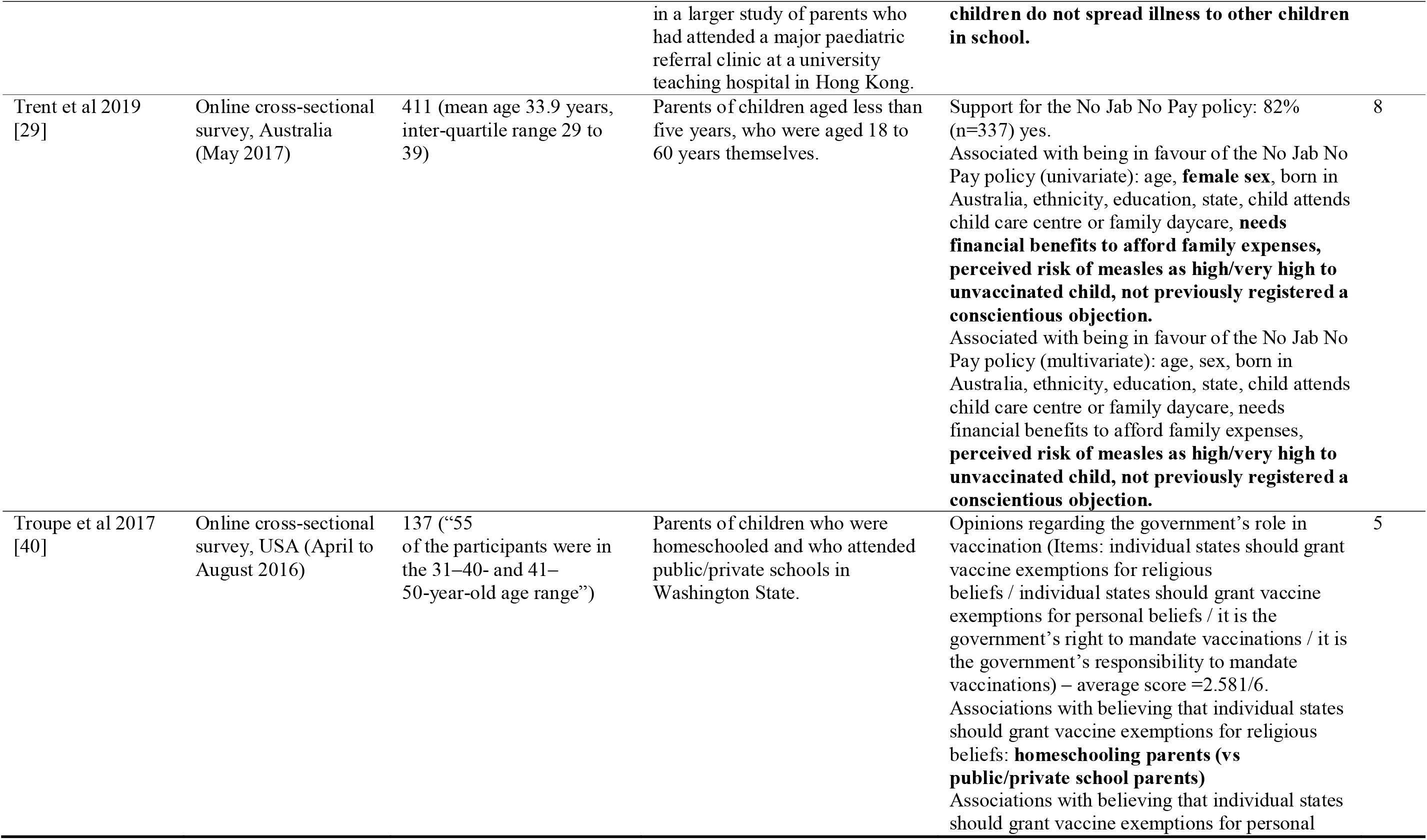
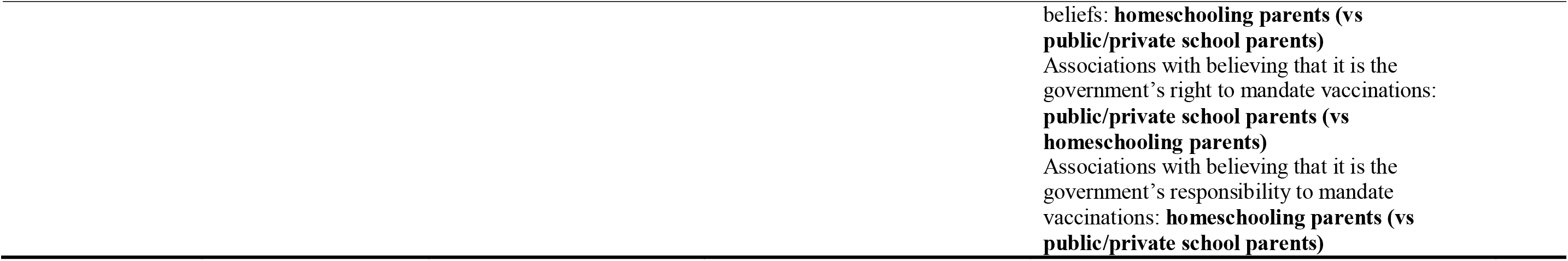

